# Natural history of epilepsy in argininosuccinic aciduria provides new insights into pathophysiology

**DOI:** 10.1101/2022.10.19.22281191

**Authors:** Nour Elkhateeb, Giorgia Olivieri, Barbara Siri, Karolina M. Stepien, Reena Sharma, Andrew Morris, Thomas Hartley, Laura Crowther, Stephanie Grunewald, Maureen Cleary, Helen Mundy, Anupam Chakrapani, Robin Lachmann, Elaine Murphy, Saikat Santra, Mari-Liis Uudelepp, Mildrid Yeo, Alicia Chan, Philippa Mills, Debora Ridout, Paul Gissen, Carlo Dionisi-Vici, Julien Baruteau

## Abstract

**Introduction:** Argininosuccinate lyase is integral to the urea cycle, which enables nitrogen waste and biosynthesis of arginine, a precursor of nitric oxide. Inherited argininosuccinate lyase deficiency causes argininosuccinic aciduria, the second most common urea cycle defect and an inherited model of systemic nitric oxide deficiency. Patients present with developmental delay, epilepsy and movement disorder. Here we aim to characterise epilepsy, a common and neurodebilitating complication in argininosuccinic aciduria.

**Patients and Methods:** We conducted a retrospective study in seven tertiary metabolic centres in the UK, Italy and Canada from 2020 to 2022 to assess the phenotype of epilepsy in ASA and correlate it with clinical, biochemical, radiological and electroencephalographic data.

**Results:** Thirty-seven patients aged 1 to 31 years old were included. Twenty-two (60%) patients presented epilepsy. Median age at epilepsy-onset was 24 months. Generalized tonic clonic and focal seizures were most common in early-onset patients whilst atypical absences were predominant in late-onset patients. Seventeen patients (77%) required antiseizure medications and 6 (27%) had partially controlled or refractory epilepsy. Epileptic patients presented with a severe neurodebilitating disease with higher rates of speech delay (*p*=0.04) and autism spectrum disorders (*p*=0.01) and more frequent arginine supplementation (*p*=0.01) compared to non-epileptic patients. Neonatal seizures were not associated with a higher risk of developing epilepsy. Biomarkers of ureagenesis did not differ between epileptic and non-epileptic patients. Epilepsy-onset in early infancy (*p*=0.05) and electroencephalographic background asymmetry (*p*=0.0007) were significant predictors of partially controlled or refractory epilepsy.

**Conclusions:** Epilepsy in argininosuccinic aciduria is frequent, polymorphic, associated with more frequent neurodevelopmental complications. We identified prognostic factors for pharmacoresistance in epilepsy. This study does not support defective ureagenesis as prominent in the pathophysiology of epilepsy but suggests roles of arginine toxicity and central dopamine deficiency.

**Key Points:**

- Epilepsy in ASA is frequent, polymorphic, occurring in early childhood and associated with a more severe neurodevelopmental phenotype.
- Early-onset epilepsy and electroencephalographic background asymmetry are prognostic for pharmaco-resistance of epilepsy in ASA.
- Hyperammonaemia is suggested not to be the primary pathophysiological mechanism for epileptogenesis in ASA.
- Central dopamine deficiency is suggested to have a role in pathophysiology of epilepsy in ASA.
- Arginine-related neurotoxicity is suggested to be associated with increased in frequency and severity of epilepsy in ASA.

## Introduction

Argininosuccinate lyase (ASL) is the only enzyme in mammals enabling endogenous arginine synthesis (1). This cytosolic enzyme, which breaks down argininosuccinic acid into arginine and fumarate, is integral to the citrulline-nitric oxide (NO) cycle, which enables NO synthesis from arginine, and the urea cycle, a liver-based pathway enabling nitrogen wasting through clearance of neurotoxic ammonia (1). ASL deficiency causes argininosuccinic aciduria (ASA) (OMIM#207900), an autosomal recessive metabolic disease and the second most common urea cycle disorder with a prevalence of 1/110,000 live birth (2). Patients present acute hyperammonaemia either in the neonatal period defined as early-onset phenotype, or later in life in late-onset presentation (3). Most patients will present a systemic phenotype with chronic neurological, hepatic, gastrointestinal, hypokalaemia and high blood pressure (4). The neurological phenotype entails intellectual and motor disability, behavioural changes, epilepsy, which can occur in absence of hyperammonaemia. The pathophysiological role of hyperammonaemia, argininosuccinate toxicity (5), and deficiency of arginine and downstream metabolites (i.e., creatine and polyamines) (2) has been proposed to account for the neurological phenotype in ASA. However novel pathophysiological insights have highlighted the role of reversible neuronal nitro-oxidative stress (6) and central catecholamine deficiency caused by NO deficiency (7,8). Standard of care in ASA relies on ammonia control using protein-restricted diet, ammonia scavengers and arginine supplementation, (9,10) with an increasing number of patients treated by liver transplantation (11).

Epilepsy is a common complication in ASA, which is thought to affect 40% of patients (4). The notion that seizures are a consequence of acute hyperammonaemia episodes is not supported by the evidence that epileptic patients usually have well-controlled ammonia levels (4,12). Recently, a mouse model of ASL-deficiency showed increased firing rate in ASL-deficient dopaminergic neurons and a lower epileptic threshold, the latter corrected by NO supplementation, highlighting the role of central catecholamine biosynthesis and its regulation by NO in the epileptogenesis of ASA (7).

Herein, we present an international multicentre retrospective study assessing the phenotype of epilepsy in ASA, its severity and correlation with age of onset of the disease, biochemical and electroencephalographic data. We show that the seizures in ASA are frequent, polymorphic and can be severe. We describe an increased rate of neurodevelopmental complications in epileptic patients, and we identify prognostic markers of epileptic severity in this population. Age at onset of hyperammonaemia, related biomarkers and therapies are not predictors for epilepsy onset and severity. Our study does not support defective ureagenesis having a prominent role in the pathophysiology of epilepsy but suggests a potential neurotoxic role of arginine supplementation and supports the pathophysiological role of central dopamine deficiency.

## Patients and Methods

We conducted a retrospective study in seven paediatric and adult tertiary metabolic centres in the UK, Italy, and Canada. Epidemiological, clinical, biochemical, radiological, and electroencephalographic (EEG) data of ASA patients with neurological disease and/or epilepsy were collected between July 2020 and June 2022.

Early-onset ASA was defined as hyperammonaemia occurring on/before 28 days of age, and late-onset ASA after 28 days of age.

Epilepsy was defined according to the International League Against Epilepsy (ILAE) 2014 guidelines (13) by any of the following conditions: 1) At least two unprovoked (or reflex) seizures occurring >24 hours apart; 2) One unprovoked (or reflex) seizure and a probability of further seizures similar to the general recurrence risk (at least 60%) after two unprovoked seizures, occurring over the next 10 years 3) Diagnosis of an epilepsy syndrome.

Complete control was defined as complete cessation of seizures after treatment, and partial control was defined as a decrease of 50% or more in seizure frequency but without a complete cessation of seizures after treatment. Failure of seizure control was defined as a less than 50% decrease in seizure frequency after treatment.

Biochemical data included the mean values of plasma ammonia, glutamine, arginine and argininosuccinic acid during compensated metabolic state. Biochemical results during the initial hyperammonaemic decompensation and after liver transplantation were not included.

Radiological data included CT/MRI brain. Electroencephalographic data included data from standard EEG or EEG telemetry. Treatment data included natural protein intake, ammonia scavengers and epilepsy treatment.

Visual related EEG abnormalities included photo-paroxysmal responses (PRR) on photic stimulation (an activation procedure), eye closure sensitivity (which describes transient epileptic abnormalities following eye closure and represents the physiological loco-regional differentiation and maturation of brain electrical activity) and scotosensitive discharge elicited by darkness.

Epilepsy severity was assessed by US Department of Veterans Affairs (VA) Seizure type and frequency rating scale (as revised for VA-2) (14), Chalfont Seizure Severity Scale (15), modified grand total EEG (16) and 2HELPS2B scores (17).

### Statistical analysis

Statistical analysis was performed using Prism 9.0 software (San Diego, CA, USA). Differences between groups were assessed using a two-tailed Mann-Whitney U test for quantitative variables and a two-tailed Fisher’s exact test for categorical data. *p* values ≤ 0.05 were considered statistically significant.

## Results

Demographics and clinical features are summarized in Tables 1 and 2. Biochemical features are summarized in Table 2 with an exhaustive dataset in Supplementary Table 1.

**Table 1:** Epidemiological and clinical characteristics of recruited ASA patients. Comparison of patients with and without epilepsy and with early- and late-onset ASA.

|  | Early onset ASA with<br>epilepsy (no=14) | Late onset ASA with<br>epilepsy (no=8) | p value * | ASA with epilepsy<br>(no=22) | ASA without epilepsy<br>(no=15) | p value † | Study population<br>(no=37) |
| --- | --- | --- | --- | --- | --- | --- | --- |
| 1. Demographic data: |  |  |  |  |  |  |  |
| Male no (%) | 10 (71.4%) | 5 (62.6%) | 0.19 | 15 (68.2%) | 6 (40%) | 0.11 | 21 (56.8%) |
| Current age (months) Median (range) | 196.5 (86-371) | 130 (42-336) | 0.5 | 192 (42-371) | 134 (15-289) | 0.4 | 144 (15-371) |
| 2. Clinical data: |  |  |  |  |  |  |  |
| Early onset ASA no (%) | 14 (100%) | 0 | N/A | 14 (63.64%) | 13 (86.7%) | 0.2 | 25 (67.6%) |
| Age at onset of symptoms of ASA (months) Median (range) | 0.2 (0.03-0.5) | 5.6 (2-12) | 0.0007 | 0.3 (0.03-12) | 0.2 (0.07-24) | 0.4 | 0.3 (0.03-24) |
| Age at Diagnosis of ASA (months) Median (range) | 0.3 (0.05-9) | 15 (3-216) | 0.0005 | 0.5 (0.05-216) | 0.25 (0.03-59) | 0.1 | 0.5 (0.03-216) |
| 3. Clinical characteristics of epilepsy |  |  |  |  |  |  |  |
| Age at onset of epilepsy (months) Median (range) | 36 (9-191.7) | 13.5 (3.5-9) | 0.06 | 24 (3.5-191.7) | N/A | N/A | N/A |
| Neonatal seizures n (%) | 7 (50%) | 0 | 0.02 | 7 (31.8%) | 2 (13.3%) | 0.3 | 9 (24.3%) |
| Seizure type n (%) |  |  |  |  |  |  |  |
| Generalized tonic clonic | 11 (78.6%) | 4 (50%) | 0.3 | 15 (68.2%) | N/A | N/A | N/A |
| Focal | 9 (64.3%) | 4 (50%) | 0.7 | 13 (59.1%) | N/A | N/A | N/A |
| Atypical Absence | 6 (42.9%) | 7 (87.5%) | 0.07 | 13 (59.1%) | N/A | N/A | N/A |
| Myoclonic & myoclonic astatic | 0 | 4 (50%) | 0.0096 | 4 (18.2%) | N/A | N/A | N/A |
| Epileptic spasms ‡ | 0 | 1 (12.5%) | 0.4 | 1 (4.6%) | N/A | N/A | N/A |
| Tonic | 0 | 1 (12.5%) | 0.4 | 1 (4.6%) | N/A | N/A | N/A |
| Multiple seizure types n (%) | 11 (78.6%) | 7 (87.5%) | 1 | 18 (81.8%) | N/A | N/A | N/A |
| Status epilepticus no (%) | 4 (28.6%) | 2 (25%) | 1 | 6 (27.3%) | N/A | N/A | N/A |
| Age at first episode of status epilepticus (months) Median (range) | 36 (9-140) | 58.5 (14-103) | 0.4 | 36 (9-140) | N/A | N/A | N/A |
| 4. Epilepsy treatment data |  |  |  |  |  |  |  |
| Treatment with ASMs n (%) | 10 (71.4%) | 7 (87.5%) | 0.6 | 17 (77.3%) | N/A | N/A | N/A |
| Treatment with ASMs n (%) (current) | 9 (64.3%) | 6 (75%) | 1 | 15 (68.2%) | N/A | N/A | N/A |
| Seizure control no (%) |  |  |  |  |  |  |  |
| Complete control | 6 (42.9%) | 3 (37.5%) | 0.7 | 9 (40.9%) | N/A | N/A | N/A |
| Partial control | 3 (21.4%) | 2 (25%) |  | 5 (22.7%) | N/A |  | N/A |
| failure of seizure control | 0 | 1 (12.5%) |  | 1 (4.5%) | N/A |  | N/A |
| Not on ASMs | 5 (35.7%) | 2 (25%) |  | 7 (31.8%) | N/A |  | N/A |
| VNS | 0 | 1 (12.5%) |  | 1 (4.6%) | N/A |  | N/A |
| 5. EEG characteristics no (%) |  |  |  |  |  |  |  |
| Visual related EEG abnormalities | 4/12 (33.3%) | 3/7 (42.9%) | 1 | 7/19 (36.8%) | N/A | N/A | N/A |
| 6. Epilepsy & EEG severity scores Mean (SD) |  |  |  |  |  |  |  |
| VA-2 seizure severity score | 33 (32.5) | 1904.2 (4936.4) | 0.4 | 713.5 (3110) | N/A | N/A | N/A |
| Chalfont Seizure Severity Scale | 56.4 (28.3) | 68 (82.3) | 0.4 | 60.6 (54.8) | N/A | N/A | N/A |
| Modified grand total EEG score (0-54) | 11.34 (5) | 15.1 (6.26) | 0.5 | 12.7 (5.8) | N/A | N/A | N/A |
| 2HELPS2B score (0-7) | 2.1 (1-4) | 2 (2-3.2) | 0.9 | 2 (1-4) | N/A | N/A | N/A |

ASA: argininosuccinic aciduria. ASM: antiseizure medication. SD: standard deviation
\* p value represents the comparison between early and late ASA patients with epilepsy † p value represents the comparison between patients with epilepsy and without
‡ This patient has a paternally inherited CACNA1A variant of uncertain significance (VUS) which could be contributing to the epilepsy phenotype. Seizures preceded the onset of initial ASA decompensation.

**Table 2:**
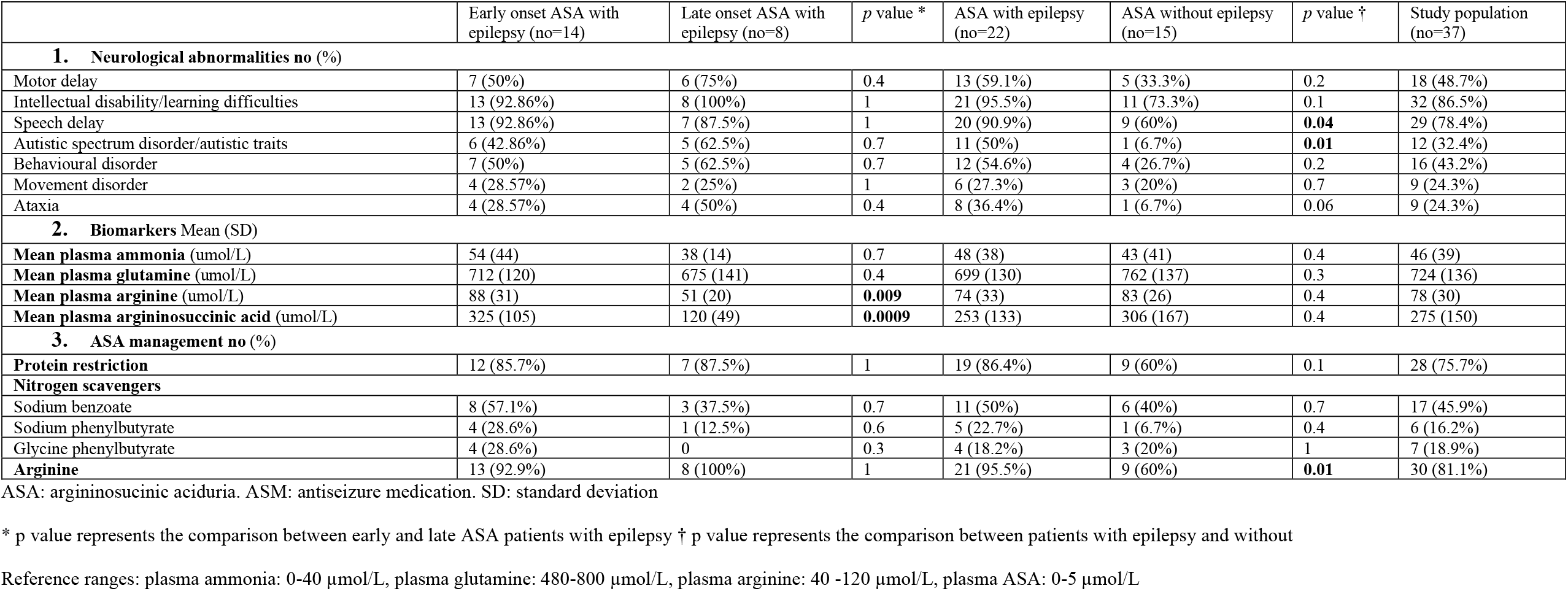
Clinical and biochemical characteristics of recruited ASA patients. Comparison of patients with and without epilepsy and with early- and late-onset ASA.

|  | Early onset ASA with epilepsy (no=14) | Late onset ASA with epilepsy (no=8) | *p* value * | ASA with epilepsy (no=22) | ASA without epilepsy (no=15) | *p* value † | Study population (no=37) |
| --- | --- | --- | --- | --- | --- | --- | --- |
| **1. Neurological abnormalities no (%)** |  |  |  |  |  |  |  |
| Motor delay | 7 (50%) | 6 (75%) | 0.4 | 13 (59.1%) | 5 (33.3%) | 0.2 | 18 (48.7%) |
| Intellectual disability/learning difficulties | 13 (92.86%) | 8 (100%) | 1 | 21 (95.5%) | 11 (73.3%) | 0.1 | 32 (86.5%) |
| Speech delay | 13 (92.86%) | 7 (87.5%) | 1 | 20 (90.9%) | 9 (60%) | **0.04** | 29 (78.4%) |
| Autistic spectrum disorder/autistic traits | 6 (42.86%) | 5 (62.5%) | 0.7 | 11 (50%) | 1 (6.7%) | **0.01** | 12 (32.4%) |
| Behavioural disorder | 7 (50%) | 5 (62.5%) | 0.7 | 12 (54.6%) | 4 (26.7%) | 0.2 | 16 (43.2%) |
| Movement disorder | 4 (28.57%) | 2 (25%) | 1 | 6 (27.3%) | 3 (20%) | 0.7 | 9 (24.3%) |
| Ataxia | 4 (28.57%) | 4 (50%) | 0.4 | 8 (36.4%) | 1 (6.7%) | 0.06 | 9 (24.3%) |
| **2. Biomarkers Mean (SD)** |  |  |  |  |  |  |  |
| Mean plasma ammonia (umol/L) | 54 (44) | 38 (14) | 0.7 | 48 (38) | 43 (41) | 0.4 | 46 (39) |
| Mean plasma glutamine (umol/L) | 712 (120) | 675 (141) | 0.4 | 699 (130) | 762 (137) | 0.3 | 724 (136) |
| Mean plasma arginine (umol/L) | 88 (31) | 51 (20) | **0.009** | 74 (33) | 83 (26) | 0.4 | 78 (30) |
| Mean plasma argininosuccinic acid (umol/L) | 325 (105) | 120 (49) | **0.0009** | 253 (133) | 306 (167) | 0.4 | 275 (150) |
| **3. ASA management no (%)** |  |  |  |  |  |  |  |
| Protein restriction | 12 (85.7%) | 7 (87.5%) | 1 | 19 (86.4%) | 9 (60%) | 0.1 | 28 (75.7%) |
| **Nitrogen scavengers** |  |  |  |  |  |  |  |
| Sodium benzoate | 8 (57.1%) | 3 (37.5%) | 0.7 | 11 (50%) | 6 (40%) | 0.7 | 17 (45.9%) |
| Sodium phenylbutyrate | 4 (28.6%) | 1 (12.5%) | 0.6 | 5 (22.7%) | 1 (6.7%) | 0.4 | 6 (16.2%) |
| Glycine phenylbutyrate | 4 (28.6%) | 0 | 0.3 | 4 (18.2%) | 3 (20%) | 1 | 7 (18.9%) |
| Arginine | 13 (92.9%) | 8 (100%) | 1 | 21 (95.5%) | 9 (60%) | **0.01** | 30 (81.1%) |

ASA: argininosuccinic aciduria. ASM: antiseizure medication. SD: standard deviation
\* *p* value represents the comparison between early and late ASA patients with epilepsy † *p* value represents the comparison between patients with epilepsy and without
Reference ranges: plasma ammonia: 0-40 µmol/L, plasma glutamine: 480-800 µmol/L, plasma arginine: 40 -120 µmol/L, plasma ASA: 0-5 µmol/L

### Demographic and Clinical characteristics

Thirty-seven patients were included with a median age of 12 years (range: 15 months-31 years) and a sex ratio male/female of 21/16. Twenty-five patients (67%) had early-onset ASA. The median age at onset of ASA symptoms was 10 days (range: 1 day-24 months). The median age at diagnosis was 15 days (range 1 day-18 years) (Table 1). Diagnosis of ASA was obtained biochemically and was confirmed genetically in 7 patients.

### Clinical characteristics of epilepsy

This study included 22 (60%) ASA patients with epilepsy, with a median age of 16 years (range: 3.5 years-31 years) and a sex ratio male/female of 15/7. Fourteen patients (64%) had early-onset ASA. The median age at onset of ASA symptoms was 10 days (range: 1 day-12 months) while the median age at diagnosis of ASA was 15 days (range: 1 day-18 years). The median age at onset of epilepsy was 24 months (range: 3.5 months-16 years). Seven patients (32%) with epilepsy had symptomatic seizures during neonatal hyperammonaemia (Table 1).

Seizure types were highly variable. The most common types were generalized tonic clonic (n=15, 68%), focal (n=13, 59%) seizures and atypical absences (n=13, 59%). Multiple seizure types were commonly present (n=18, 82%). These were either concurrent or evolving from one type to another. One patient presented with epileptic spasms before the age of 1 year old that subsequently evolved to atypical absence seizures. Six patients (27%) presented status epilepticus. Median age at onset of the first episode of status epilepticus was 36 months (range 9-12 years). The main trigger of status epilepticus was febrile intercurrent illnesses with associated hyperammonaemic decompensation (Table 1).

Seventeen patients (77%) required anti-seizure medications (ASMs). ASMs were tapered off and stopped completely in two patients. The number of ASMs currently required was 1 (n=10; 59%), 2 (n=4; 24%) or 3 (n=1; 6%). One patient required vagal nerve stimulation for epilepsy control, which was initiated in late childhood, with initial favourable response for 2 years followed by recurrence of refractory seizures. ASMs enabled complete seizure control in 9 patients (60%), partial seizure control in 5 patients (23%) while seizures were pharmaco-resistant in one patient (Table 1).

### Additional neurological features in ASA epileptic patients

Twenty-one patients (95%) presented with variable degrees of intellectual disability and learning difficulties. Speech delay was seen in 20 patients (91%), motor delay in 13 patients (59%) and behavioural disorders in 12 patients (55%). Autistic features were seen in 11 patients (50 %) and a formal diagnosis of autistic spectrum disorder was made in 9 patients (41%). Ataxia was seen in 8 patients (36%) while movement disorders were seen in 6 patients (27%) (Table 2).

### Neuroimaging characteristics in ASA epileptic patients

Brain MRI was performed in 13 patients (59%). Neuroimaging abnormalities were seen in 8/13 (62%). The most common abnormalities included cerebral white matter changes (n=4), cerebral atrophy (n=2) and basal ganglia abnormalities (n=2). Multiple bilateral hemisphere infarcts crossing arterial territories (n=1), cerebellar atrophy (n=1). Neuroimaging was normal in 5 (38%) patients (Supplementary table 1).

### Electroencephalographic characteristics in ASA epileptic patients

Electroencephalography was performed in 19 patients (86%). Generalized background slowing was seen in 12 patients (63%). Focal background slowing was seen in 7 patients (32%), predominantly in temporal (n=4) and occipital (n=3) regions. Background asymmetry was noted in 8 patients (42%). Inter-ictal epileptogenic activity was detected in 15 patients (79%) with focal discharges (n=13, 68%), generalized discharges (n=4, 21%), or paroxysmal response to hyperventilation (n=1, 5%). Visual-related EEG abnormalities (n=6, 32%) was observed with photo-paroxysmal responses on photic stimulation with flickering light (n=2, 11%) between 6-60 Hz (n=1, 5%) and 16-30 Hz (n=1, 5%), eye closure sensitivity (n=3, 16%), and scotosensitive discharges in darkness (n=1, 5%) (Table 1; Supplementary table 1). There was no fixation-off sensitivity. Hypsarrhythmia pattern with epileptic spasms was seen in one patient (5%). This patient had a paternally inherited CACNA1A variant of uncertain significance (VUS). Focal epileptic discharges were central (n=7, 37%), temporal (n=5, 26%), frontal (n=4, 21%), occipital (n=1, 5%), parietal (n=2, 11%). Discharges were unilateral (n=3, 16%) or bilateral/multifocal (n=9, 47%). Unilateral electrical status epilepticus during slow-wave sleep (ESES) was observed in 1 patient. Age at photo-paroxysmal responses ranged between 1 to 18 years while age at eye closure sensitivity ranged between 5.5 to 10 years (Supplementary table 1).

### Epilepsy and EEG severity scores

Epilepsy clinical severity scores were calculated for all ASA epileptic patients (n=22). The mean values of clinical severity scores were 713.5 for the VA-2 seizure severity score (range: 7.5-14964) and 61 for the Chalfont Seizure Severity Scale (range: 10-268). EEG severity scores were calculated for 18 patients. The mean values of EEG severity scores were 12.66/54 (range: 4-27) for the mean Modified grand total EEG score and 2.33/7 (range: 1-4) for the 2HELPS2B score (Table 1).

### Non-neurological characteristics in ASA epileptic patients

Transaminitis was the most common feature (n=11, 50%), followed by hepatomegaly (n=11, 50%). Tubulopathy with hypokalaemia were seen in 6 patients (27%) and hair changes as trichorrhexis nodosa were seen in 5 patients (23%) (Supplementary table 1).

### Biochemical characteristics in ASA epileptic patients

Mean plasma ammonia after the neonatal period/initial metabolic decompensation was 46 *μ*mol/L (range: 14.5-190 *μ*mol/L, reference range 0-40 µmol/L). Mean plasma glutamine was 724 *μ*mol/L (range: 481-1060 *μ*mol/L, reference range 480-800 µmol/L) and mean plasma arginine was 78 *μ*mol/L (range: 25-139 *μ*mol/L, reference range 40 -120 µmol/L). Mean plasma argininosuccinate was 275 *μ*mol/L (range: 17-641, reference range 0-5 µmol/L) (Table 2).

### Metabolic management of ASA epileptic patients

Nineteen patients (86%) were on protein restricted diet. Eleven patients (50%) were on sodium benzoate with a median dose of 188 mg/kg/day. Five patients (23%) were on sodium phenylbutyrate with a median dose of 230 mg/kg/day. Four patients (18%) were on glycerol phenylbutyrate with a median dose of 189 mg/kg/day. Five patients (23%) were on two nitrogen scavengers, 10 patients (45%) were on one scavenger and 7 patients (32%) did not require any nitrogen scavenger. Twenty-one patients (95%) were on arginine supplementation with a median dose of 124 mg/kg/day (Supplementary table 1). Liver transplantation was performed in six patients and infusion of hepatic stem cells (as a part of a clinical trial) was performed in one patient. The six transplanted patients had stopped arginine and ammonia scavenger therapy and liberalized the dietary protein intake.

### Comparison between ASA patients with and without epilepsy

We compared data from 22 epileptic and 15 non-epileptic ASA patients (Tables 1, 2 and supplementary Table 1).). Neurological and behavioural abnormalities were significantly more frequent in epileptic patients, especially for speech delay (*p*=0.04) and autistic spectrum disorder (*p*=0.01) and with a trend for ataxia symptoms (*p*=0.06). 7/9 (78%) of patients with symptomatic seizures in the neonatal period developed epilepsy. Neonatal seizures were not associated with a higher risk of developing epilepsy (*p*=0.3), (Relative risk at age 5 years of age = 0.96). No statistical difference was observed for plasma ammonia, glutamine, arginine and argininosuccinic acid. Epileptic patients were more frequently treated with arginine supplementation (*p*=0.01). They received higher mean dose of sodium benzoate (182 vs 152 mg/kg/day; *p*=0.5) and glycerol phenylbutyrate (180 vs 124 mg/kg/day; *p*=0.2). No significant difference was observed for the following variables: gender, age of disease-onset, neonatal seizures, neuroimaging features and non-neurological abnormalities and ammonia scavenger requirement.

### Comparison between early- and late-onset ASA epileptic patients

We compared the early- and late-onset cohorts of ASA epileptic patients with 14 and 8 patients respectively (Tables 1, 2 and supplementary Table 1). Both groups showed variable seizure types with generalised tonic clonic seizures and focal seizures being the predominant types in early-onset ASA patients while atypical absence seizures were the predominant type in late-onset ASA patients. Early-onset ASA patients had significantly higher mean plasma arginine (*p*=0.009) and argininosuccinic acid (*p*=0.0009). Mean plasma ammonia and glutamine were non significantly different. There was a trend for the patients in early-onset group to be on more ammonia scavengers and a higher dose of sodium benzoate than the late-onset group. The numbers of patients with protein restriction and oral arginine supplementation were not different between groups. No significant difference was observed for the following variables: gender, age at epilepsy-onset, number of AEDs, seizure control, epilepsy severity scores, additional neurological features, non-neurological features, EEG characteristics, EEG severity scores, neuroimaging abnormality.

### Comparison of ASA epileptic patients with well-controlled versus partially controlled or refractory epilepsy

We compared ASA patients with well-controlled epilepsy (n=16, 73%) versus ASA patients with partially controlled (n=5, 23%) or refractory to ASM (n=1, 4 %). Well-controlled epileptic patients were either without or with ASMs (Table 3 and supplementary table 2). Partially controlled and refractory epilepsy was significantly associated with epilepsy-onset at a younger age (median 36 months versus 13.5 months; *p*=0.05), EEG background asymmetry (*p*=0.0007) and did not present with visual-sensitive EEG abnormalities although this last finding was not significant (*p*=0.15). No significant difference was observed for the following variables: gender, age of disease-onset, neonatal seizures, seizure types, epilepsy and EEG severity scores, neuroimaging abnormalities, neurological and non-neurological abnormalities, biochemical biomarkers, ammonia scavengers and arginine dose and requirement.

**Table 3:** Comparison between ASA patients with well-controlled versus partially controlled or refractory epilepsy.

|  | Well-controlled epilepsy (n=16) | Partially controlled and refractory epilepsy (n=6) | *p* value |
| --- | --- | --- | --- |
| **1. Demographic data:** |  |  |  |
| **Sex n (%)** |  |  |  |
| Male | 12 (75%) | 3 (50%) | 0.3 |
| Current age (months) Median (range) | 196.5 (42-371) | 168 (116-336) | 0.7 |
| **2. Clinical data:** |  |  |  |
| **Onset of ASA** |  |  |  |
| Early onset ASA | 11 (68.8%) | 3 (50%) | 0.6 |
| Age at onset of symptoms (months) Median (range) | 0.3 (0.03-12) | 0.5 (0.07-3.5) | 0.7 |
| Age at Diagnosis (months) Median (range) | 0.3 (0.05-216) | 3.3 (0.3-12) | 0.3 |
| **3. Clinical characteristics of epilepsy** |  |  |  |
| Age at onset of epilepsy (months) Median (range) | 36 (7-191.7) | 13.5 (3.5-60) | **0.05** |
| Neonatal seizures n (%) | 5 (31.3%) | 2 (33.3%) | 1 |
| **4. EEG characteristics no (%)** |  |  |  |
| Background asymmetry | 2/14 (14.3%) | 6/6 (100%) | **0.0007** |
| **5. Epilepsy & EEG severity scores Mean (SD)** |  |  |  |
| VA-2 seizure severity score (mean) | 35.2 (37.8) | 2522.2 (5564.3) | 0.1 |
| Chalfont Seizure Severity Scale (mean) | 50.9 (34) | 86.5 (83.7) | 0.5 |
| Modified grand total EEG score (mean) (0-54) | 11.97 (5) | 14.9 (7.4) | 0.4 |
| 2HELPS2B score (mean) (0-7) | 2.4 (0.8) | 2 (0.7) | 0.3 |
| **6. Biomarkers mean (SD)** |  |  |  |
| Mean plasma ammonia ($\mu\text{mol/L}$) | 49 (43.3) | 45.6 (10.8) | 0.3 |
| Mean plasma glutamine ($\mu\text{mol/L}$) | 719.5 (131.3) | 644.9 (107.9) | 0.3 |
| Mean plasma arginine ($\mu\text{mol/L}$) | 78.3 (30.6) | 63.9 (35.2) | 0.3 |
| Mean plasma argininosuccinic acid ($\mu\text{mol/L}$) | 275.7 (138.1) | 164.5 (43.5) | 0.2 |

ASA: argininosuccinic aciduria, EEG: electroencephalogram, SD: standard deviation
Reference ranges: plasma ammonia: 0-40 $\mu\text{mol/L}$ , plasma glutamine: 480-800 $\mu\text{mol/L}$ , plasma arginine: 40 -120 $\mu\text{mol/L}$ , plasma ASA: 0-5 $\mu\text{mol/L}$

### Correlation between severity scores and biological markers in ASA epileptic patients

Number of ASMs showed a significant negative correlation with plasma arginine levels in ASA epileptic patients (Spearmann’s correlation coefficient rs= -0.47, *p*= 0.03). Chalfont seizure severity scale showed a significant positive correlation with plasma arginine levels in ASA epileptic patients (Spearmann’s correlation coefficient rs= 0.43, *p*= 0.047). Number of ASMs and other epilepsy and EEG severity scores did not show any other significant correlation with selected biomarkers (plasma ammonia, glutamine, argininosuccinate, arginine) (Supplementary table 3).

## Discussion

Epilepsy is a common feature in urea cycle disorders affecting 3-14% of patients (18,19). ASA is a systemic disease with complex pathophysiology and a model of inherited NO deficiency (20). This international multicentric retrospective study is the largest to focus on the epilepsy phenotype in ASA patients. Our work suggests even higher frequency (60%) of epilepsy in ASA patients, compared with previous publications reporting incidence of 42% (4) to 55% (12). Seizure types are polymorphic, although our study shows that tonic clonic and multifocal seizures are more frequently observed in early-onset ASA whilst atypical absences are the more common seizure type in late-onset ASA. The epilepsy phenotype occurs early in the natural history of ASA with a median at 24 months of age, which is earlier than the median ages of 3 years and 5.5 years reported by Grioni *et al* (12) and Baruteau *et al* (4), respectively. The epilepsy phenotype is severe with 27% of patients presenting partial control or refractory seizures, and 27% presenting with status epilepticus, usually triggered by febrile illness. The main prognostic factors predicting epilepsy pharmaco-resistance were epilepsy-onset in early infancy before the age of 2 years and electroencephalographic background asymmetry. The absence of visual-related EEG abnormalities (i.e., photosensitivity or eye closure sensitivity on EEG) could be a pejorative indicator, although this association was not significant in this work.

ASA phenotype is associated with a severe neurodebilitating disease, with characterised by developmental delay, learning difficulties, ataxia, and motor symptoms (4). Our study shows that epileptic ASA patients have a significantly higher rate of speech delay, autistic spectrum disorder and a trend for ataxia symptoms.

Acute symptomatic seizures and subclinical electrographic seizures are observed in 33-50% of neonatal hyperammonaemia (21-23). Acute symptomatic seizures develop during the rise of glutamine levels, which occurs before ammonia increase (24). Our study showed that neonatal hyperammonaemic seizures in ASA were not associated with a higher risk of developing epilepsy.

The prevalence of epilepsy is also common in another urea cycle disorder, arginase deficiency (OMIM 207800), affecting 60%-75% of patients. It has been suggested that the pathophysiology of epilepsy is caused by high arginine, subsequent downstream metabolite guanidinoacetate and its neurotoxicity (25-27). Guanidinoacetate metabolites have a neurotoxic role by impairing redox homeostasis and mitochondrial bioenergetics (28,29). In ASA, increased guanidinoacetate has been documented by MRI spectroscopy, (4,30,31) and could play a role in the pathophysiology of epilepsy (32). In this work, arginine supplementation was significantly more frequent in the epilepsy group, although plasma arginine levels were not different. Plasma arginine levels had a positive correlation with epilepsy severity as indicated by Chalfont seizure severity scale. This suggests that exogenous arginine supplementation may have a role in the epileptogenesis in ASA by raising neurotoxic guanidinoacetate levels, as suggested by previous reports (2) (12). Additional clinical data on arginine metabolites including guanidinoacetate levels in cerebrospinal fluid would be of interest in ASA patients to better characterise this association. Interestingly high dose of arginine supplementation has shown liver toxicity in this disorder (9).

Although argininosuccinate is neurotoxic at high dose generating oxidative stress (5), no significant difference in plasma argininosuccinate levels was observed between epileptic and non-epileptic patients, or well- and partially controlled patients. Plasma levels do not reflect adequately the central effect of toxic metabolites produced *in situ*. Argininosuccinic acid is likely trapped in the brain like other dicarboxylic or tricarboxylic acids (33,34), therefore plasma levels are not reliable markers to assess cerebrospinal fluid levels.

No significant difference in age of onset of hyperammonaemia, ureagenesis biomarkers and number and dose of ammonia scavengers was observed between epileptic and non-epileptic, well-controlled versus partially controlled or refractory patients. Higher plasma argininosuccinate and scavenger doses in early-onset patients support previous reports (4). These findings suggest that hyperammonaemia is not the primary pathophysiological mechanism for epileptogenesis (2,4). Recently Lerner *et al* have shown a link between epileptogenesis in ASA and central catecholamine deficiency (7). A novel mouse model with ASL-deficiency exclusively in dopaminergic neurons *Asl*^*flox/flox*^*;TH Cre*^*+/-*^ showed that ASL-deficient dopaminergic neurons from the locus coeruleus have abnormal electrophysiology *i*.*e*. more frequent potential firing activity and a sharper after-hyperpolarisation recovery slope (7). This was associated with higher sensitivity to medication-induced epilepsy, which was restored by supplementation with NO donor (7). The antiepileptic role of the locus coeruleus in limiting the spread and the duration of epilepsy (35,36) is well recognised and is essential for efficacious vagal nerve stimulation (37,38). One of the patients in this work showed only a short and transient response to VNS, which could be explained by defective catecholamine synthesis in the locus coeruleus. Additionally, the locus coeruleus plays a central role in regulating arousal, wakefulness, and the sleep-wake pattern (39). This work highlighted a high frequency of visual-related EEG abnormalities (32%), triggered by photic stimulation, eye closure and elimination of retinal light stimulation. Interestingly a deficit in dopaminergic inhibitory neurotransmission has been described in generalised photosensitive epilepsy (40) and epileptic photosensitivity in progressive myoclonus epilepsies (41). This further supports the association between photosensitive-related EEG abnormalities and dopamine deficiency in ASA.

This work has limitations due to the small number of patients affected by this rare disease and retrospective analysis. Our findings warrant further prospective studies from larger cohorts of patients, which could be achieved via registries of patients affected by urea cycle disorders (42). Understanding the neurological phenotype from levels of metabolites measured in plasma is inadequate. Analysing metabolites from cerebrospinal fluid and MRI spectroscopy with prospective monitoring will provide better tools to understand the pathophysiology of the epilepsy and neurological disease in ASA. Developing alternatives models with induced pluripotent stem cells derived neurons or neuronal or organotypic cultures will provide surrogates to better study the complex pathophysiology of this disorder (43,44).

## Conclusion

Epilepsy is a cardinal symptom of the encephalopathy observed in ASA. Epilepsy is frequent, polymorphic, occurring in early childhood and associated with a more severe neurodevelopmental phenotype. Early-onset of epilepsy before the age of 2 years and electroencephalographic background asymmetry are prognostic markers for pharmaco-resistance of epilepsy. Age at onset of first hyperammonaemia and ureagenesis biomarkers and therapies do not differ between epileptic and non-epileptic patients or well and poorly controlled epileptic patients, suggesting that hyperammonaemia is not the primary pathophysiological mechanism for epileptogenesis in ASA. Arginine supplementation was significantly more frequent in epileptic patients and plasma arginine levels had a positive correlation with epilepsy severity, in favour of arginine-related neurotoxicity. Poor response to vagal nerve stimulation in one patient and high frequency of visual-related EEG abnormalities provide clinical context and support the pathophysiological role of central dopamine deficiency.

## Data Availability

The authors confirm that the data supporting the findings of this study are available within the article [and/or] its supplementary materials or available upon request by contacting the senior author.

## Declarations

All authors reviewed and approved the final version of the manuscript.

### Details of funding

This study was supported by the United Kingdom Medical Research Council Clinician Scientist Fellowship MR/T008024/1 (to JB) and NIHR Great Ormond Street Hospital Biomedical Research Centre (to JB). The views expressed are those of the author(s) and not necessarily those of the NHS, the NIHR or the Department of Health.

### Conflicts of interest/Competing interests

None

### Data Sharing and Data Availability

The data that supports the findings of this study are available in the supplementary material of this article.

### Availability of data and material

The authors confirm that the data supporting the findings of this study are available within the article [and/or] its supplementary materials.

## Ethics approval

### Consent to participate

This research study was conducted retrospectively from medical notes. Participants’ data were recorded anonymously. Informed consent approved by the National Research Ethics Service Committee London-Bloomsbury (13/LO/0168) was obtained from all participants and/or legal guardians for the following centres: Great Ormond Street Hospital for Children NHS Trust, National Hospital for Neurology and Neurosurgery, Evelina London Children’s Hospital, Salford Royal NHS Foundation Trust and Manchester Centre for Genomic Medicine. The centres, Birmingham Children’s Hospital, Bambino Gesù Children’s Hospital in Rome and University of Alberta, did not require consents from their institutional review board due to the collection of anonymous data.

### Consent for publication

This research study was conducted retrospectively from medical notes. Participants’ data were recorded anonymously. Informed consent was obtained from all participants and/or legal guardians for the following centres: Great Ormond Street Hospital for Children NHS Trust, National Hospital for Neurology and Neurosurgery, Evelina London Children’s Hospital, Salford Royal NHS Foundation Trust,and Manchester Centre for Genomic Medicine. The centres, Birmingham Children’s Hospital, Bambino Gesù Children’s Hospital in Rome and University of Alberta, did not require consents from their institutional review board due to the collection of anonymous data.

## Acknowledgments

We thank Dr Marios Kaliakatsos, Consultant Paediatric Neurologist and Dr Stewart Boyd, Consultant Neurophysiologist, at Great Ormond Street Hospital NHS Trust, London, UK for their valuable comments on EEG abnormalities.

---

**Supplementary table 1:**
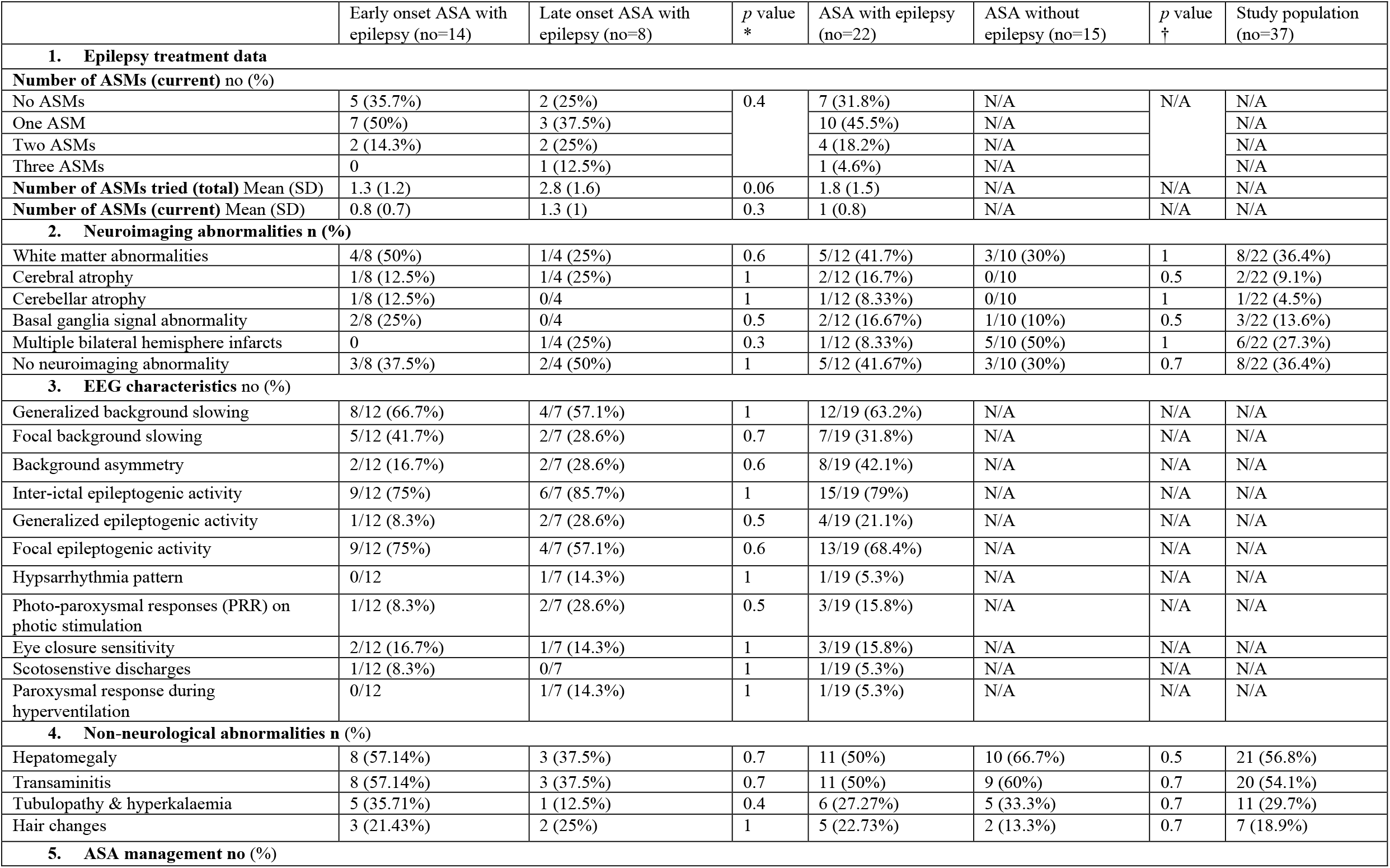
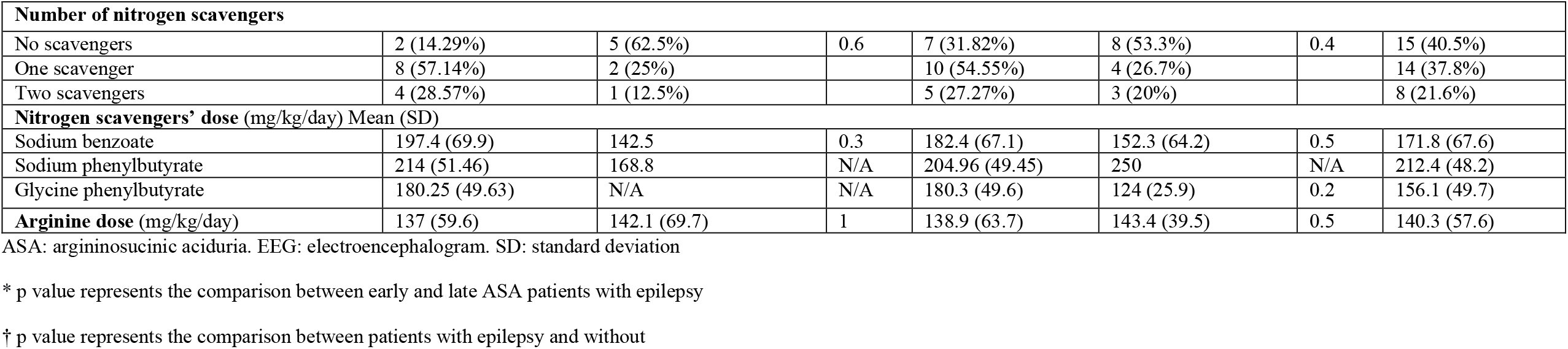
Epilepsy and ASA treatment, neuroimaging, EEG and non-neurological characteristics of ASA patients with epilepsy and comparison data between early and late onset ASA groups with epilepsy.

**Supplementary table 2:** Comparison between ASA patients with well-controlled versus partially controlled or refractory epilepsy.

|  | Well-controlled epilepsy (n=16) | Partially controlled and refractory epilepsy (n=6) | p value |
| --- | --- | --- | --- |
| 1. Clinical characteristics of epilepsy |  |  |  |
| Seizure type n (%) |  |  |  |
| Generalized tonic clonic | 10 (62.5%) | 5 (83.3%) | 0.6 |
| Focal | 9 (56.3%) | 4 (66.7%) | 1 |
| Atypical Absence | 8 (50%) | 5 (83.3%) | 0.3 |
| Myoclonic and myoclonic astatic | 3 (18.8%) | 2 (33.3%) | 0.6 |
| Epileptic spasms * | 1 (6.3%) | 0 | 1 |
| Tonic | 0 | 1 (16.7%) | 0.3 |
| Multiple seizure types n (%) | 12 (75%) | 6 (100%) | 0.5 |
| Status epilepticus n (%) | 3 (18.8%) | 3 (50%) | 0.3 |
| 2. EEG characteristics no (%) |  |  |  |
| Generalized background slowing | 8/14 (57.1%) | 4/6 (66.7%) | 1 |
| Focal background slowing | 6/14 (42.9%) | 1/6 (16.7%) | 0.4 |
| Inter-ictal epileptogenic activity | 12/14 (85.7%) | 3/6 (50%) | 0.1 |
| Generalized epileptogenic activity | 4/14 (28.6%) | 0/6 | 0.3 |
| Focal epileptogenic activity | 11/14 (78.6%) | 2/6 (33.3%) | 0.1 |
| Photo-paroxysmal responses (PRR) on photic stimulation | 3/14 (21.4%) | 0/6 | 0.5 |
| Eye closure sensitivity | 3/14 (21.4%) | 0/6 | 0.5 |
| 3. Neurological abnormalities n (%) |  |  |  |
| Motor delay | 9 (56.3%) | 4 (66.7%) | 1 |
| Intellectual disability/learning difficulties | 15 (93.8%) | 6 (100%) | 1 |
| Speech delay | 15 (93.8%) | 5 (83.3%) | 0.5 |
| Autistic spectrum disorder/autistic traits | 6 (37.5%) | 5 (83.3%) | 0.1 |
| Behavioural disorder | 8 (50%) | 4 (66.7%) | 0.6 |
| Movement disorder | 4 (25%) | 2 (33.3%) | 1 |
| Ataxia | 4 (25%) | 4 (66.7%) | 0.1 |
| Abnormal neuroimaging | 6/10 (60%) | 2/3 (66.7%) | 1 |
| 4. Non-neurological abnormalities no (%) |  |  |  |
| Hepatomegaly | 10 (62.5%) | 1 (16.7%) | 0.1 |
| Transaminitis | 8 (50%) | 3 (50%) | 1 |
| Tubulopathy & hyperkalemia | 5 (31.3%) | 1 (16.7%) | 0.6 |
| Hair changes | 3 (18.8%) | 2 (33.3%) | 0.6 |
| 5. ASA management |  |  |  |
| Protein restriction n (%) | 13 (81.3%) | 6 (100%) | 0.5 |
| Nitrogen scavengers n (%) |  |  |  |
| Sodium benzoate | 7 (43.8%) | 4 (66.7%) | 0.6 |
| Sodium phenylbutyrate | 5 (31.3%) | 0 | 0.3 |
| Glycine phenylbutyrate | 4 (25%) | 0 | 0.5 |
| Arginine n (%) | 15 (93.8%) | 6 (100%) | 1 |
| No of nitrogen scavengers no (%) |  |  |  |
| No scavengers | 6 (37.5%) | 1 (16.7%) | 0.5 |
| One scavenger | 6 (37.5%) | 4 (66.7%) |  |
| Two scavengers | 4 (25%) | 1 (16.7%) |  |
| Nitrogen scavengers' dose (mg/kg/day) Mean (SD) |  |  |  |
| Sodium benzoate | 208.8 (52.3) | 150.72 (69.16) | 0.2 |
| Sodium phenylbutyrate | 214 (51.5) | 168.8 | N/A |
| Glycine phenylbutyrate | 180.3 (49.6) | N/A | N/A |
| Arginine | 144.2 (64.2) | 125.8 (60.5) | 0.6 |

ASA: argininosuccinic aciduria, EEG: electroencephalogram, SD: standard deviation
\* This patient has a paternally inherited CANCA1 variant of uncertain significance (VUS) which could be contributing to the epilepsy phenotype. Seizures preceded the onset of initial ASA decompensation.

**Supplementary table 3:** Correlation between clinical features and biomarkers in ASA patients with epilepsy.

|  | Plasma ammonia | Plasma glutamine | Plasma arginine | Plasma argininosuccinic acid |
| --- | --- | --- | --- | --- |
| Number of ASMs (current) | 0.2 | 1 | **0.03** | 0.2 |
| VA-2 seizure severity score | 0.9 | 0.3 | 0.5 | 0.3 |
| Chalfont seizure severity scale | 0.3 | 0.7 | **0.047** | 0.4 |
| Modified grand total EEG score | 0.8 | 0.4 | 0.9 | 1 |
| 2HELPS2B score | 0.2 | 0.1 | 0.6 | 0.7 |

ASM: antiseizure medication.

## Notes

### Competing Interest Statement

The authors have declared no competing interest.

### Author Declarations

This research study was performed under the ethical oversight from the National Research Ethics Committee London-Bloomsbury (study reference identifier 13/LO/0168). Informed consent was obtained from all participants and/or legal guardians for Great Ormond Street Hospital for Children, National Hospital for Neurology and Neurosurgery, Evelina London Children's Hospital, Salford Royal NHS Foundation Trust and Manchester Centre for Genomic Medicine. Anonymous data collection did not require consents for the following centres: Birmingham Children's Hospital, Bambino Gesu Children's Hospital in Rome and University of Alberta.

## References

1. Erez A, Nagamani SC, Lee B. Argininosuccinate lyase deficiency-argininosuccinic aciduria and beyond. Am J Med Genet C Semin Med Genet. 2011;157C:45–53. doi:10.1002/ajmg.c.30289

2. Baruteau J, Diez-Fernandez C, Lerner S, et al. Argininosuccinic aciduria: Recent pathophysiological insights and therapeutic prospects. J Inherit Metab Dis. 2019;42:1147–1161. doi:10.1002/jimd.12047

3. Tuchman M, Lee B, Lichter-Konecki U, et al. Cross-sectional multicenter study of patients with urea cycle disorders in the United States. Mol Genet Metab. 2008;94:397–402. doi:10.1016/j.ymgme.2008.05.004

4. Baruteau J, Jameson E, Morris AA, et al. Expanding the phenotype in argininosuccinic aciduria: need for new therapies. J Inherit Metab Dis. 2017;40:357–368. doi:10.1007/s10545-017-0022-x

5. Seminotti B, da Silva JC, Ribeiro RT, Leipnitz G, Wajner M. Free Radical Scavengers Prevent Argininosuccinic Acid-Induced Oxidative Stress in the Brain of Developing Rats: a New Adjuvant Therapy for Argininosuccinate Lyase Deficiency?. Mol Neurobiol. 2020;57:1233–1244. doi:10.1007/s12035-019-01825-0

6. Baruteau J, Perocheau DP, Hanley J, et al. Argininosuccinic aciduria fosters neuronal nitrosative stress reversed by Asl gene transfer. Nat Commun. 2018;9:3505. doi:10.1038/s41467-018-05972-1

7. Lerner S, Anderzhanova E, Verbitsky S, et al. ASL Metabolically Regulates Tyrosine Hydroxylase in the Nucleus Locus Coeruleus. Cell Rep. 2019;29:2144-2153.e7. doi:10.1016/j.celrep.2019.10.043

8. Lerner S, Eilam R, Adler L, et al. ASL expression in ALDH1A1+ neurons in the substantia nigra metabolically contributes to neurodegenerative phenotype. Hum Genet. 2021;140:1471–1485. doi:10.1007/s00439-021-02345-5

9. Nagamani SC, Lee B, Erez A. Optimizing therapy for argininosuccinic aciduria. Mol Genet Metab. 2012;107:10–14. doi:10.1016/j.ymgme.2012.07.009

10. Nagamani SCS, Erez A, Lee B. Argininosuccinate Lyase Deficiency. 2011 Feb 3 [Updated 2019 Mar 28]. In: Adam MP, Everman DB, Mirzaa GM, et al., editors. GeneReviews® [Internet]. Seattle (WA): University of Washington, Seattle; 1993-2022. Available from: https://www.ncbi.nlm.nih.gov/books/NBK51784/

11. Molema F, Martinelli D, Hörster F, et al. Liver and/or kidney transplantation in amino and organic acid-related inborn errors of metabolism: An overview on European data. J Inherit Metab Dis. 2021;44(3):593–605. doi:10.1002/jimd.12318

12. Grioni D, Furlan F, Corbetta C, et al. Epilepsy and argininosuccinic aciduria. Neuropediatrics. 2011;42:97–103. doi:10.1055/s-0031-1280795

13. Fisher RS, Acevedo C, Arzimanoglou A, et al. ILAE official report: a practical clinical definition of epilepsy. Epilepsia. 2014;55:475–482. doi:10.1111/epi.12550

14. Cramer JA, French J. Quantitative assessment of seizure severity for clinical trials: a review of approaches to seizure components. Epilepsia. 2001;42:119–129. doi:10.1046/j.1528-1157.2001.19400.x.

15. Duncan JS, Sander JW. The Chalfont Seizure Severity Scale. J Neurol Neurosurg Psychiatry. 1991;54:873–876. doi:10.1136/jnnp.54.10.873

16. Barcelon EA, Mukaino T, Yokoyama J, et al. Grand Total EEG Score Can Differentiate Parkinson’s Disease From Parkinson-Related Disorders. Front Neurol. 2019;10:398. doi:10.3389/fneur.2019.00398

17. Struck AF, Ustun B, Ruiz AR, et al. Association of an Electroencephalography-Based Risk Score with Seizure Probability in Hospitalized Patients. JAMA Neurol. 2017;74:1419–1424. doi:10.1001/jamaneurol.2017.2459

18. Kölker S, Valayannopoulos V, Burlina AB, et al. The phenotypic spectrum of organic acidurias and urea cycle disorders. Part 2: the evolving clinical phenotype [published correction appears in J Inherit Metab Dis. 2015 No;38(6):1157–8. Garcia Cazorla, Angeles [corrected to Garcia-Cazorla, Angeles]]. J Inherit Metab Dis. 2015;38(6):1059-1074. doi:10.1007/s10545-015-9840-x

19. Toquet S, Spodenkiewicz M, Douillard C, et al. Adult-onset diagnosis of urea cycle disorders: Results of a French cohort of 71 patients. J Inherit Metab Dis. 2021;44:1199–1214. doi:10.1002/jimd.12403

20. Kho J, Tian X, Wong WT, et al. Argininosuccinate Lyase Deficiency Causes an Endothelial-Dependent Form of Hypertension. Am J Hum Genet. 2018;103:276–287. doi:10.1016/j.ajhg.2018.07.008

21. Campistol J. Epilepsy in Inborn Errors of Metabolism with Therapeutic Options. Semin Pediatr Neurol. 2016;23:321–331. doi:10.1016/j.spen.2016.11.006

22. Pearl PL. Amenable Treatable Severe Pediatric Epilepsies. Semin Pediatr Neurol. 2016;23:158–166. doi:10.1016/j.spen.2016.06.004

23. Ah Mew N, Simpson KL, Gropman AL, et al. Urea Cycle Disorders Overview. 2003 Apr 29 [Updated 2017 Jun 22]. In: Adam MP, Mirzaa GM, Pagon RA, et al., editors. GeneReviews® [Internet]. Seattle (WA): University of Washington, Seattle; 1993-2022. Available from: https://www.ncbi.nlm.nih.gov/books/NBK1217/

24. Wiwattanadittakul N, Prust M, Gaillard WD, et al. The utility of EEG monitoring in neonates with hyperammonemia due to inborn errors of metabolism. Mol Genet Metab. 2018;125:235–240. doi:10.1016/j.ymgme.2018.08.011

25. Deignan JL, De Deyn PP, Cederbaum SD, et al. Guanidino compound levels in blood, cerebrospinal fluid, and post-mortem brain material of patients with argininemia. Mol Genet Metab. 2010;100 Suppl 1:S31–S36. doi:10.1016/j.ymgme.2010.01.012

26. Huemer M, Carvalho DR, Brum JM, et al. Clinical phenotype, biochemical profile, and treatment in 19 patients with arginase 1 deficiency. J Inherit Metab Dis. 2016;39:331–340. doi:10.1007/s10545-016-9928-y

27. Chandra SR, Christopher R, Ramanujam CN, Harikrishna GV. Hyperargininemia Experiences over Last 7 Years from a Tertiary Care Center. J Pediatr Neurosci. 2019;14:2–6. doi:10.4103/jpn.JPN_1_19

28. Kolling J, Wyse AT. Creatine prevents the inhibition of energy metabolism and lipid peroxidation in rats subjected to GAA administration. Metab Brain Dis. 2010;25:331–338. doi:10.1007/s11011-010-9215-9

29. Marques EP, Ferreira FS, Santos TM, et al. Cross-talk between guanidinoacetate neurotoxicity, memory and possible neuroprotective role of creatine. Biochim Biophys Acta Mol Basis Dis. 2019;1865:165529. doi:10.1016/j.bbadis.2019.08.005

30. van Spronsen FJ, Reijngoud DJ, Verhoeven NM, Soorani-Lunsing RJ, Jakobs C, Sijens PE. High cerebral guanidinoacetate and variable creatine concentrations in argininosuccinate synthetase and lyase deficiency: implications for treatment?. Mol Genet Metab. 2006;89:274–276. doi:10.1016/j.ymgme.2006.02.005

31. Sijens PE, Reijngoud DJ, Soorani-Lunsing RJ, Oudkerk M, van Spronsen FJ. Cerebral 1H MR spectroscopy showing elevation of brain guanidinoacetate in argininosuccinate lyase deficiency. Mol Genet Metab. 2006;88:100–102. doi:10.1016/j.ymgme.2005.10.013

32. Schulze A, Ebinger F, Rating D, Mayatepek E. Improving treatment of guanidinoacetate methyltransferase deficiency: reduction of guanidinoacetic acid in body fluids by arginine restriction and ornithine supplementation. Mol Genet Metab. 2001;74:413–419. doi:10.1006/mgme.2001.3257

33. Tamai I, Tsuji A. Transporter-mediated permeation of drugs across the blood-brain barrier. J Pharm Sci. 2000;89:1371–1388. doi:10.1002/1520-6017(200011)89:11<1371::aid-jps1>3.0.co;2-d

34. Kölker S, Sauer SW, Surtees RA, Leonard JV. The aetiology of neurological complications of organic acidaemias--a role for the blood-brain barrier. J Inherit Metab Dis. 2006;29:701–706. doi:10.1007/s10545-006-0415-8

35. Szot P, Weinshenker D, White SS, et al. Norepinephrine-deficient mice have increased susceptibility to seizure-inducing stimuli. J Neurosci. 1999;19:10985–10992. doi:10.1523/JNEUROSCI.19-24-10985.1999

36. Giorgi FS, Ferrucci M, Lazzeri G, et al. A damage to locus coeruleus neurons converts sporadic seizures into self-sustaining limbic status epilepticus. Eur J Neurosci. 2003;17:2593–2601. doi:10.1046/j.1460-9568.2003.02692.x

37. Krahl SE, Clark KB, Smith DC, Browning RA. Locus coeruleus lesions suppress the seizure-attenuating effects of vagus nerve stimulation. Epilepsia. 1998;39:709–714. doi:10.1111/j.1528-1157.1998.tb01155.x

38. Fornai F, Ruffoli R, Giorgi FS, Paparelli A. The role of locus coeruleus in the antiepileptic activity induced by vagus nerve stimulation. Eur J Neurosci. 2011;33:2169–2178. doi:10.1111/j.1460-9568.2011.07707.x

39. Daneault V, Dumont M, Massé É, Vandewalle G, Carrier J. Light-sensitive brain pathways and aging. J Physiol Anthropol. 2016;35:9. Published 2016 Mar 15. doi:10.1186/s40101-016-0091-9

40. Quesney LF, Reader TA. (1990). Role of Dopamine in Generalized Photosensitive Epilepsy: Electroencephalographic and Biochemical Aspects. In: Avoli, M., Gloor, P., Kostopoulos, G., Naquet, R. (eds) Generalized Epilepsy. Birkhäuser Boston. https://doi.org/10.1007/978-1-4684-6767-3_21

41. Mervaala E, Andermann F, Quesney LF, Krelina M. Common dopaminergic mechanism for epileptic photosensitivity in progressive myoclonus epilepsies. Neurology. 1990;40:53–56. doi:10.1212/wnl.40.1.53

42. Posset R, Garbade SF, Boy N, et al. Transatlantic combined and comparative data analysis of 1095 patients with urea cycle disorders-A successful strategy for clinical research of rare diseases. J Inherit Metab Dis. 2019;42:93–106. doi:10.1002/jimd.12031

43. Diez-Fernandez C, Hertig D, Loup M, et al. Argininosuccinate neurotoxicity and prevention by creatine in argininosuccinate lyase deficiency: An in vitro study in rat three-dimensional organotypic brain cell cultures. J Inherit Metab Dis. 2019;42:1077–1087. doi:10.1002/jimd.12090

44. Duff C, Baruteau J. Modelling urea cycle disorders using iPSCs. NPJ Regen Med. 2022;7(1):56. Published 2022 Sep 26. doi:10.1038/s41536-022-00252-5

